# Adjuvant rituximab and elevated intratumoural CD8 expression are associated with sustained disease control after radiotherapy in early-stage follicular lymphoma: TROG99.03

**DOI:** 10.1101/2024.08.09.24311704

**Authors:** Michael P. MacManus, John F Seymour, Hennes Tsang, Richard Fisher, Colm Keane, Muhammed B Sabdia, Soi C Law, Jay Gunawardana, Karthik Nath, Stephen H Kazakoff, Mario L Marques-Piubelli, Daniela E Duenas, Michael R Green, Daniel Roos, Peter O’Brien, Andrew McCann, Richard Tsang, Sidney Davis, David Christie, Chan Cheah, Benhur Amanuel, Tara Cochrane, Jason Butler, Anna Johnston, Mohamed Shanavas, Li Li, Claire Vajdic, Robert Kridel, Victoria Shelton, Samantha Hershenfield, Tara Baetz, David Lebrun, Nathalie Johnson, Marianne Brodtkorb, Maja Ludvigsen, Francesco d’Amore, Ella R Thompson, Piers Blombery, Maher K Gandhi, Joshua WD Tobin

## Abstract

**Background:** We report extended follow-up of TROG99.03, a randomised phase III trial in early-stage follicular lymphoma (ESFL) including new information on the role of adjuvant rituximab and translational studies.

**Methods:** Patients with ESFL were randomised to involved-field radiotherapy (IFRT) or IFRT plus 6-cycles cyclophosphamide/vincristine/prednisolone (IFRT+CVP). From 2006 rituximab was added to IFRT+CVP (IFRT+R-CVP). Clinical and multi-omic parameters were evaluated. Findings were validated in two independent ESFL cohorts (99 and 60 patients respectively).

**Findings:** Between 2000-2012, 150 (75 per arm) patients were recruited. 48% were positron emission tomography (PET)-staged. Per protocol, at median follow-up 11.3-years, progression-free survival (PFS) remained superior for IFRT+(R)CVP vs. IFRT (hazard ratio [HR]=0.60, 95%CI=0.37-0.98, p=0.043; 10-year PFS 62% vs. 43%) respectively. Although no significant difference in overall survival was observed (HR=0.44, 95%CI=0.16-1.18, p=0.11, 10-year OS 95% vs 84%), patients receiving IFRT+(R)CVP experienced fewer composite (histological transformation and death) events (p=0.045). PFS of IFRT+R-CVP-treated patients compared with all other treatments lacking rituximab (IFRT alone plus IFRT+CVP) was superior (HR=0.36, 95%CI=0.13-0.82, p=0.013). Amongst PET-staged patients, PFS differences between IFRT+R-CVP vs. IFRT were maintained (HR=0.38, 95%CI=0.16-0.89, p=0.027) indicating benefit distinct from stage migration. FL-related mutations and *BCL2*-translocations were not associated with PFS. However, by multivariate analysis elevated CD8A gene expression in diagnostic biopsy tissue was independently associated with improved PFS (HR=0.45, 95%CI=0.26-0.79, p=0.037), a finding confirmed in both ESFL validation cohorts. CD8A gene expression was raised (p=0.02) and CD8+ T-cell density higher within follicles in ESFL vs. advanced-stage FL (p=0.047). Human leucocyte antigen class I specific neoantigens were detected in 43% of patients, suggesting neoantigen-specific CD8+ T-cells have a role in confining the spread of the disease.

**Interpretation:** Adjuvant R-CVP and elevated intratumoural CD8 expression were independently associated with sustained disease control after radiotherapy in ESFL.

**Funding:** Cancer Council Victora; National Health and Medical Research Council; Leukaemia Foundation; Mater Foundation.

## Introduction

Follicular lymphoma (FL) is the most common indolent Non-Hodgkin lymphoma in adults, with an annual incidence of 2.5-3.3 per 100 000 population.(1, 2, 3) Approximately 20% of patients with FL present with localised disease (stage I-II) or ‘early-stage FL’ (ESFL). Curative-intent local radiotherapy (RT), given as involved-field (IFRT) or involved-site (ISRT) to 24-30Gy, achieves 10-year progression-free survival (PFS) of approximately 50% with low toxicity and is a widely-used standard therapy in ESFL.(4) Relapses generally occur outside irradiated volumes. Non-randomised trials suggest that the addition of anti-CD20 monoclonal antibodies (mAb) is well tolerated in ESFL patients but in contrast to stage III-IV or ‘advanced-stage FL’ (ASFL),(5, 6) no consensus exists regarding the role of systemic therapy or incorporation of anti-CD20 mAb such as rituximab as part of initial treatment for ESFL.(7, 8) The Trans-Tasman Radiation Oncology Group (TROG) study TROG99.03 represents the only modern randomised controlled trial (RCT) comparing IFRT alone with combined modality therapy comprising IFRT plus systemic therapy.(9) Patients were randomly-assigned to either IFRT alone or IFRT plus 6-cycles of cyclophosphamide, vincristine and prednisolone (IFRT+CVP). From 2006 rituximab was added to combined modality therapy (IFRT+R-CVP). Initially, reported outcomes were that progression-free survival (PFS) in the IFRT+(R)CVP arm was superior to IFRT alone, but overall survival (OS) was similar between arms.(9) The PFS curves did not separate until approximately 5 years. Data were insufficiently mature to compare long-term outcomes between patients receiving rituximab vs. those that did not.

Emerging data have identified significant biological distinctions between ESFL and ASFL. These include reduced frequency of genomic alterations including *BCL2*-translocations in ESFL vs. ASFL, and also distinct immune gene expression signatures.(10, 11, 12) Furthermore, there are fewer pro-tumoural macrophages within the tumour microenvironment (TME) in ESFL,(10) indicating that the TME of ESFL is less immunosuppressive than that of ASFL. There is also a modest but significant increased frequency of CD8+ T-cells in ESFL vs. ASFL.(10) The mechanistic basis for the increase in intratumoural CD8+ T-cells is unknown. It remains to be tested whether genetic aberrations and TME characteristics influence the outcome of patients with ESFL.

Here, with 30 months additional follow-up, we report updated clinical endpoints. This additional follow-up is sufficient to provide new information on the role of rituximab within combined modality therapy (IFRT+R-CVP) The updated trial endpoints are presented here as intention-to-treat (ITT) analyses. Translational studies were conducted to identify genomic and microenvironmental characteristics which are unique to limited-stage FL and to explore the impact of these biological features on treatment response.

## Methods

### Study design and participants

The TROG99.03 trial was conducted by the Trans-Tasman Radiation Oncology Group (TROG), with participation of Australasian Leukaemia and Lymphoma Group and Princess Margaret Cancer Centre, Toronto, Canada. Between 2000-2012, this trial recruited 150 patients with a diagnosis of grade I-IIIa, stage I/II FL with life expectancy >5-years, and adequate haematologic and renal function. Previous radiotherapy or chemotherapy, prior malignancy (excluding non-melanoma skin cancer), pregnancy, specified infectious diseases, or thoracic disease too extensive for safe IFRT, rendered patients ineligible.

Detailed trial procedures (Figure 1A and supplemental) have been previously published.(9) Briefly, after written informed consent, patients were allocated to either IFRT alone (30-36Gy) (IFRT arm) or identical IFRT followed by 6-cycles of 3-weekly ‘CVP’ (cyclophosphamide/vincristine/prednisolone) (IFRT+CVP). Randomization was stratified for centre, stage, age (≤ 59 v ≥ 60 years), and from 2006 on, positron emission tomography (PET) staging, using the minimization technique incorporating a random element. Patient characteristics were balanced between treatment arms as shown (Table S1). Reflecting evolving standards of care, in mid-2006 the protocol was amended to add rituximab (375mg/m^2^) to combined modality therapy (i.e. IFRT+R-CVP). As it became more widely adopted, ^18^F-fluorodeoxyglucose-positron-emission tomography (FDG-PET) staging was introduced as a randomisation stratification factor (but not mandated for study entry) from mid-2006. The protocol specified regular post-treatment follow-up visits and annual computed tomography surveillance imaging for at least 10-years. The trial was conducted in accordance with the Declaration of Helsinki.

**Figure 1:**
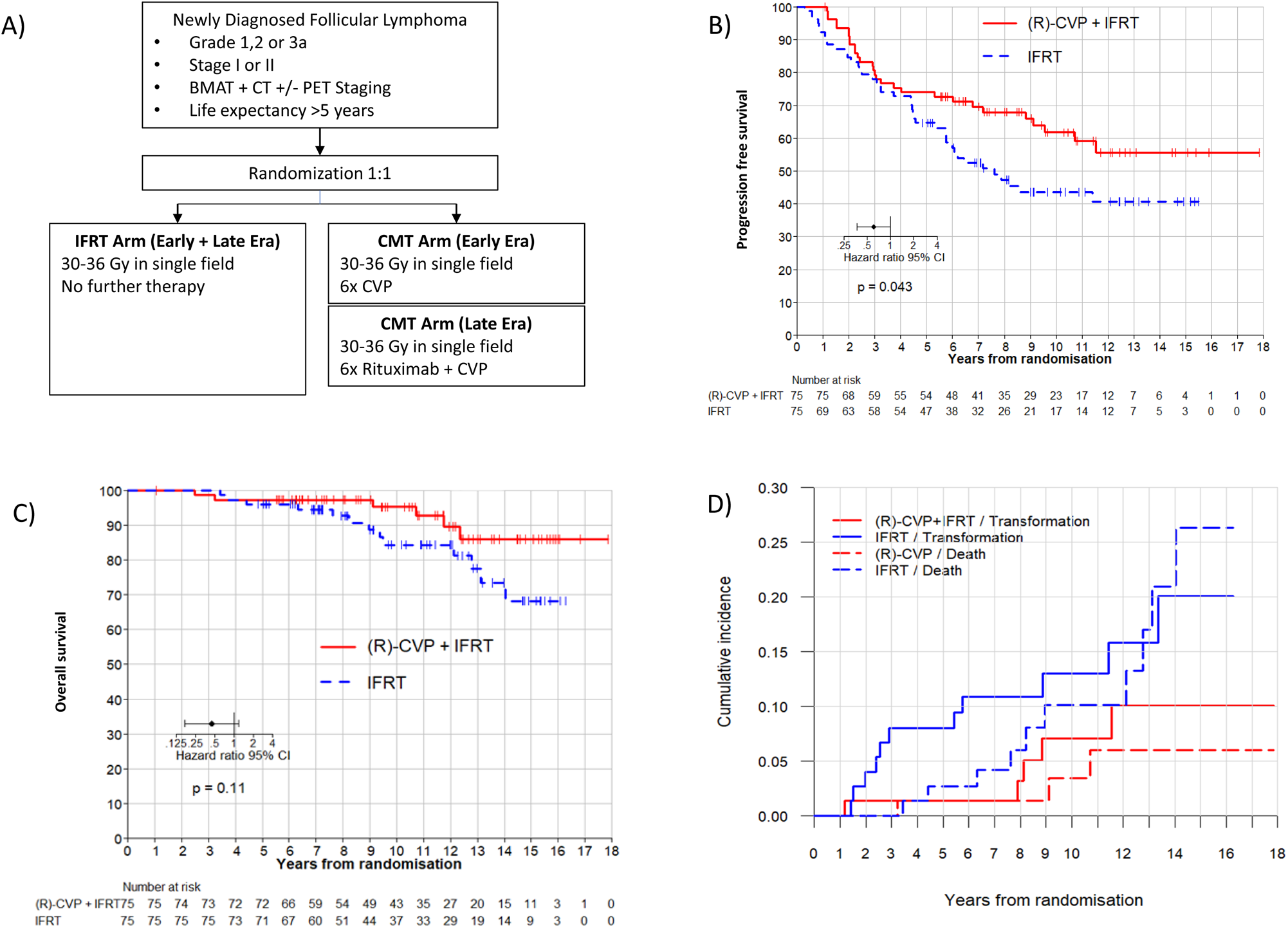
Outcomes of 150 patients with ESFL registered to the TROG99.03 trial with extended follow-up. A) Clinical trial schema outlining randomization to IFRT vs. combined modality treatment (CMT) (i.e. IFRT+(R)CVP) arms. B) Progression-free survival of IFRT vs. IFRT+(R)CVP arms. C) Overall survival of IFRT+(R)CVP vs. IFRT arms. D) Hazard curves demonstrating cumulative incidence of death and histologic transformation in IFRT+(R)CVP and IFRT treatment arms.

Correlative studies were performed on 101 available diagnostic tissue samples from the TROG99.03 study in the ‘discovery cohort’ (Figure S1). Analysis was by treatment received. Two independent, non-trial ESFL cohorts (AusESFL and CanESFL) were assembled separately (Figure S2) to validate the gene expression findings from TROG99.03. Both independent cohorts included only PET and bone marrow biopsy staged patients. AusESFL (n=99) is a multicentre retrospective cohort from six Australian and one American site between 2005-2014Patients included in this cohort were >18 years, were treatment-naïve and were treated with IFRT with or without adjuvant immunochemotherapy. CanESFL (n=60) consisted of retrospective samples from patients with stage 1 ESFL from three Canadian and two European centres treated uniformly with IFRT. All patients were aged ≥18. Table S2 compares the clinical characteristics of the discovery and validation cohorts. Exclusion criteria for both cohorts were a previous diagnosis of lymphoma, previous lymphoma therapy and those with grade IIIb FL or composite or transformed disease. Samples with insufficient RNA or DNA or failed quality control were excluded from analysis. For experiments that required comparisons between ESFL with ASFL, an additional, contemporaneous retrospective cohort of 68 patients with stage III/IV disease (termed AusASFL), drawn from the same centres as AusESFL, were tested.

### Gene expression, mutational analysis and neoantigen profiling

RNA was digitally quantified using the NanoString® 770-gene PanCancer Immune panel (730 immune-related genes plus 40 housekeeping genes) and nSolver® analysis support.(13) Sequencing used a 330 gene targeted ‘PanHaem V2.0’ panel. A minimum of 200ng of DNA was used. For Human Leucocyte Antigen (HLA)-class I typing, a computational pipeline was utilized involving sequence-based germline (blood) 6-digit genotyping. Neoantigens were filtered according to strong binding affinity to HLA-class I over wild-type, as per established ‘Tumor Neoantigen Selection Alliance’ (TESLA) consortium guidelines including a stringent threshold for HLA-class I binding affinity of <50nM.(14) Details are provided in the supplement.

### Immunohistochemistry

Paraffin-embedded tissue microarray (TMA) samples were stained with CD8 using Leica Bond RX slide stainer (Leica Biosystems) following an optimized protocol per antibody. Tonsil and appendix tissues sections were used as positive controls. Two independent pathologists, blinded to all clinical details, reviewed images for suitability for automated digital image analysis using an Aperio AT2 slide scanner (Leica Biosystems) at 20x magnification., Pathologists supervised the training for automated segmentation of the tissue into intra-follicular and extra-follicular areas. Details are provided in the supplemental.

### Statistics

For clinical trial outcomes, the prespecified primary analysis compared PFS between IFRT and (R)CVP+IFRT stratified by the minimization variables (stage, age, PET-staging) by Cox proportional hazard ratio based on intention-to-treat analysis. Exploratory biological analysis was conducted according to ‘treatment received’ analysis (expanded on in supplemental results). Categorical data were compared with Fisher’s exact or χ2 test, when appropriate, and continuous data using two-tailed paired t tests. Continuous data were compared using Wilcoxon rank-sum testing; Benjamini-Hochberg correction was used for multiple hypothesis testing where appropriate. Survival analysis was performed using R version 2023.12.1+402. For NanoString® analysis in the discovery cohort, dichotomization used optimal levels of gene expression using maximally selected rank statistics. The Kaplan-Meier method was used to estimate OS and PFS and survival curves were compared using the log-rank test. Hazard ratios (HRs) and 95% confidence intervals (95%CIs) were estimated using Cox proportional hazards regression analysis. Variables that showed different distribution across groups (p<0.1) were included in the Cox regression models that used PFS as the dependent variable to identify potential independent prognostic factors. 2-sided p-values <0.05 were considered significant.

### Role of the funding source

Funding sources had no involvement in study design, collection, analysis, and interpretation of the data, or in writing the report or decision to submit this paper for publication. PB, HT, ERT, SHK had access to the raw sequencing data. HT, MKG, MPM, JFS, RF and JWDT had access to raw clinical data of TROG99.03 patients. HT, MKG and JWDT had access to raw clinical data of validation patients. The corresponding authors had full access to all the data in the study and the final responsibility for the decision to submit for publication.

## Results

### Study Participants

Between February 2000 and July 2012, 150 patients were recruited from 21 centres in Australia, New Zealand and Canada, an accrual period of 12.5-years. The initial published analysis was performed at a closeout date of July 2015.(9) Follow-up continued to December 31^st^, 2017, to enable this updated analysis. Median follow-up time in this analysis was 11.3-years (range 4.4-17.8). For the 8 patients (5.3% of study cohort) lost to extended follow-up, median was 9.9-years. A total of 88 patients were enrolled in the pre-rituximab era and 62 following the 2006 amendment with 31/62 patients assigned to receive IRFT+R-CVP. The 101 cases included in the discovery cohort were representative of the TROG99.03 cohort (Table S2).

### Updated Outcomes from TROG99.03 Trial

An ITT analysis was performed for primary study endpoints. Adjusted for the stratification variables, (stage, PET staging and age ≥60), the primary endpoint PFS, remained significantly superior in the IFRT plus (R)-CVP arm (HR=0.60, 95%CI=0.37-0.98, p=0.043) (Figure 1B). Overall, 70 patients (29 (R)-CVP arm, 41 IFRT arm) experienced PFS events, 14 of which occurred during the extended follow-up period. Estimated 10-year PFS was 62% for the (R)-CVP arm and 43% for the IFRT arm respectively. No further disease progressions were recorded beyond 12-years in either arm.

There was no statistically significant difference in OS between arms (HR=0.44, 95%CI=0.16-1.18, p=0.11, Figure 1C). In total, 19 of 150 patients had died by the closeout date. More than twice as many deaths occurred in the IFRT arm (n=13) as in the (R)-CVP arm (n=6). The OS curves for the 2 arms largely overlapped until approximately 8-years of follow-up before diverging in the IFRT and IFRT+(R)CVP arms respectively. Histologic transformation (HT) to aggressive lymphoma also occurred in 16 patients with 11 and 5 cases occurring in the IFRT and IFRT+(R)CVP arms respectively. By last follow-up, 69 IFRT+(R)CVP arm patients were alive of whom 2 had transformed. Six IFRT+(R)CVP patients had died of whom 3 had transformed. By last follow-up 62 IFRT patients were alive of whom 8 had transformed. Thirteen IFRT patients had died of whom 3 had transformed. Patients randomised to IFRT+(R)CVP experienced significantly fewer composite events (histological transformation and deaths) than patients who received IFRT alone (p=0.045), (Figure 1D).

### The Effect of Rituximab-Containing Systemic Therapy: Intention to Treat Analysis

Extended follow-up enabled estimation of the effect of rituximab on patient outcomes compared to patients not receiving rituximab over the same period. A stratified ITT analysis, comparing 31 patients randomised to rituximab (IFRT+R-CVP) with 31 patients assigned IFRT alone in the rituximab era, demonstrated superior PFS in rituximab-randomised patients compared to patients randomised to IFRT alone (HR=0.26, 95%CI=0.084-0.82, p=0.021).

There was no significant difference in PFS between PET-staged vs. non-PET staged patients in the whole cohort (Figure 2A). Amongst PET-staged patients the difference between IFRT+(R)-CVP vs. IFRT was maintained (HR=0.38, 95%CI=0.16-0.89, p=0.027), indicating the benefit from rituximab was not attributable to stage migration (Figure 2B). The PFS was superior in patients who received rituximab compared to the patients in the same era randomised to IFRT alone (HR=0.34, 95%CI=0.12-0.99; p=0.049, Figure 2C)

**Figure 2:**
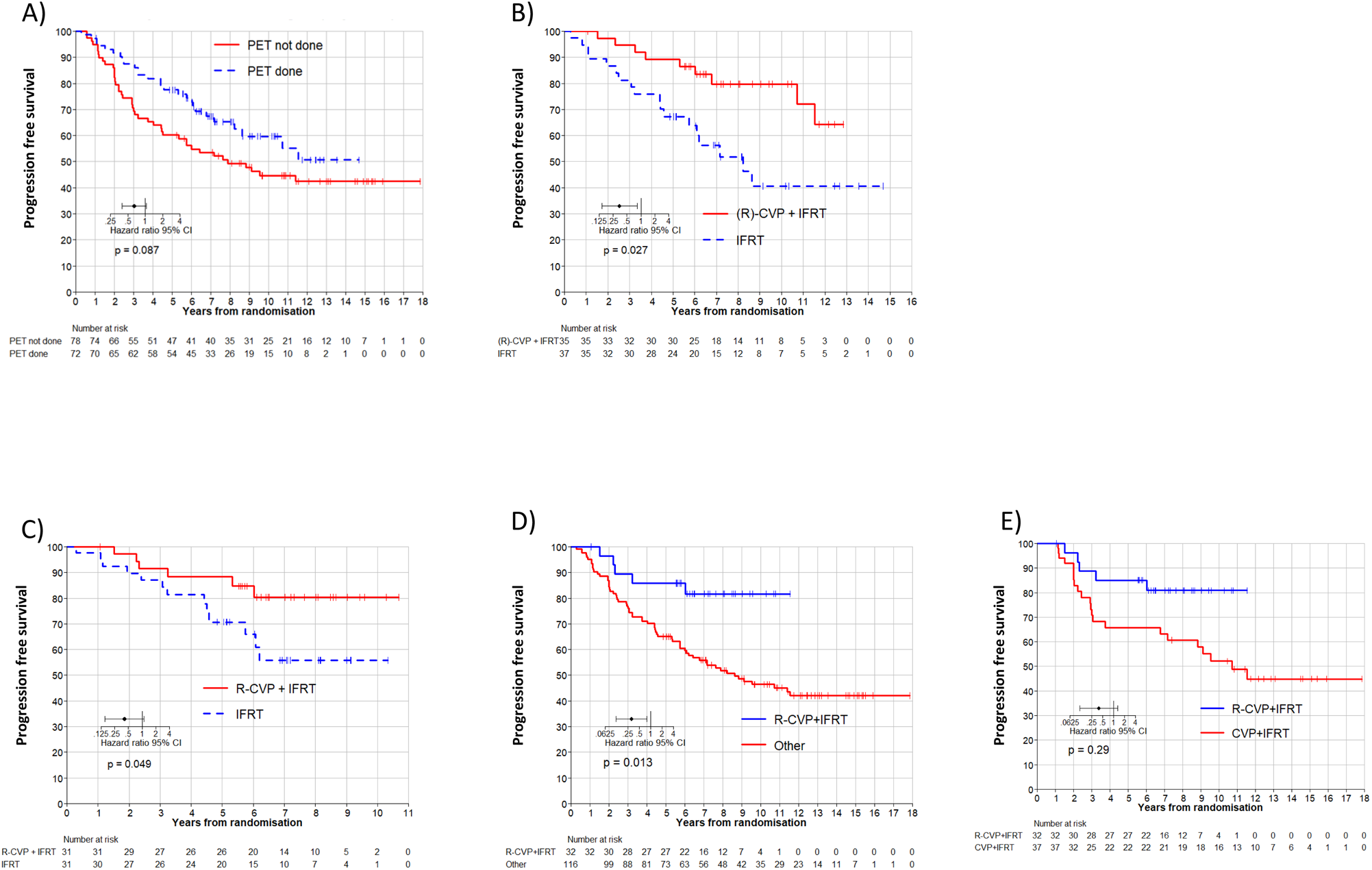
Outcomes based on baseline FDG-PET staging, period, and rituximab. A) Progression-free survival in all patients in the TROG99.03 trial based on whether baseline FEG-PET was used as a staging modality. B) Progression-free survival in the subset of TROG99.03 patients who were PET-staged. C) Progression-free survival of CMT vs IFRT in patients enrolled in the late era where rituximab was uniformly included in the CMT arm. D) Progression-free survival, per treatment received, of R-CVP treated patients compared with all non-rituximab containing treatments. E) Progression-free survival, per treatment received, of R-CVP treated patients compared with CVP treated patients.

### The Effect of Rituximab-Containing Systemic Therapy: Analysis by Treatment Received

Three patients chose to receive IFRT+R-CVP, despite being randomised either to IFRT (n=1) or IFRT+CVP (n=2). One patient randomised to IFRT+R-CVP received only IFRT. Two of the 150 patients received no IFRT, either declining all treatment or receiving CVP alone. When the PFS of all 32 patients who received IFRT+R-CVP was compared to all 116 patients who received either IFRT alone or IFRT+CVP (‘rituximab naive’), across the entire trial cohort, IFRT+R-CVP treatment was markedly superior (HR 0.36, 95%CI=0.13-0.82, p=0.013, Figure 2D). When compared across treatment era, those receiving IFRT had similar outcomes regardless of whether treated in early-or late-era after adjusting for PET-staging (HR=0.82, 95%CI=0.39-1.73; p=0.60). Analysis of patients treated on the IFRT+(R)-CVP arm by era is underpowered, however demonstrated a 10-year PFS of 81% in late-era (IFRT+R-CVP) vs 52% treated in early-era (IFRT+CVP only)although this did not reach statistical significance (HR=0.51, 95%CI=0.15-1.79; p=0.29)(Figure 2E).

### Second Malignancies

Acute and late toxicities of the trial have been previously reported and remained unchanged at this analysis. With extended follow-up, data on secondary malignancies can now be reported. Overall, the rates of secondary malignancy were similar in the two arms. Subsequent non-melanoma skin cancers were seen in 2 vs 6 patients and solid cancers (including melanoma) were seen in 11 vs 10 patients in IFRT and IFRT+(R)CVP arms respectively (Table S3). Only one therapy-related myeloid neoplasm was observed in this trial occurring in a patient who received IFRT+(R)CVP followed by multiple lines of subsequent systemic therapy after relapse.

### Clinical Prognostic Factors

Identifying high-risk ESFL patients may guide the application of IFRT+(R)CVP. Potential good-risk clinical prognostic markers (female sex, stage I, grade I-II, age ≤60-years, normal lactate dehydrogenase, normal β2-Microglobulin, presence of extranodal disease) and their univariate associations with PFS are demonstrated in Table S4. As previously reported extranodal involvement (HR=0.13, 95%CI=0.02-0.92, p=0.041) remained a favourable prognostic factor, noting that 5/12 extranodal cases were isolated duodenal involvement, which is now accepted to be particularly indolent.(15) A normal β2-Microglobulin was also associated with superior PFS (HR=0.47, 95%CI=0.22-0.99, p=0.046). On stepwise multivariable analysis of prognostic factors, only extranodal involvement (HR=0.14, 95%CI=0.02-1.01, p=0.05) approached significant association with PFS. The FLIPI score could be calculated in 93 patients (62%), classifying 88% of cases as low-risk (0–1) and 12% of patients as intermediate-risk (2) and was not significant for PFS (p=0.13).

### Genomic Prognosticators of ESFL in TROG99.03 Cohort

There were 101 TROG99.03 tissues available and of sufficient quality for genomic analysis (n=52 IFRT+(R)CVP arm, n=49 IFRT arm). Non-silent mutations were identified in 87.1% of samples with a median of 7.7 variants per patient, with as expected mutations in *CREBBP*, *KMT2D*, *TNFSRF14* and *BCL2* the most frequent. Univariate analysis for frequent mutations occurring in >10% (10 mutations) demonstrated no association with PFS after adjusting for multiple correction (Table S5).

*BCL2*-translocations occurred in 40% of cases which is slightly lower but broadly similar in frequency to another recent ESFL study (49%).(11). There was no prognostic significance of *BCL2*-translocations for PFS (HR=0.10, 95%CI=0.64-1.91, p=0.729) on univariate analysis. Mutation frequencies, stratified by *BCL2*-translocation status, were compared by Wilcoxon rank sum and corrected by Benjamini-Hochberg correction for multiple hypothesis testing. Consistent with previous reports, (10, 11) cases with *BCL2*-translocations were enriched for mutations in *CREBBP, KMT2D* and *BCL2* (68% vs. 43%, p=0.016; 65% vs. 34%, p=0.0041; 50% vs. 18%, p<0.001 respectively), whereas in contrast to those studies, STAT6 mutations did not differ (p=0.99), which may reflect the relatively low number of STAT6 mutations observed in our series (Figure 3). (11)

**Figure 3:**
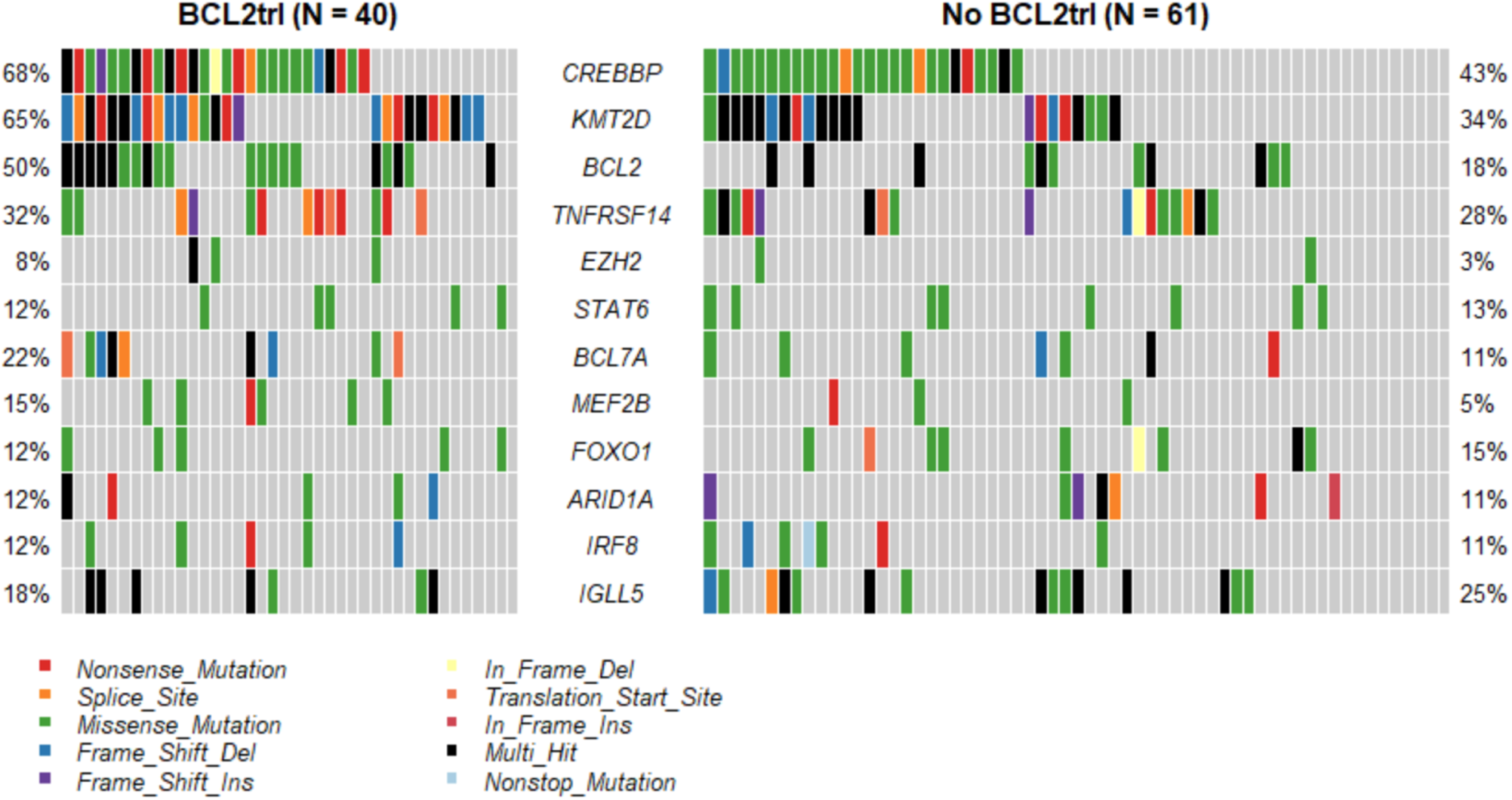
Waterfall plot demonstrating mutation profile of 101 patients with ESFL enrolled in the TROG99.03 trial. The frequencies and nature of the mutations of the top 12 genes are shown, with patients stratified by BCL2-translocation.

### Elevated intratumoural CD8A gene expression was associated with improved PFS

(16)All 97 TROG99.03 tissue samples that met RNA quality control standards underwent digital quantification. Previous studies in ASFL indicate T-cells appear protective and macrophage markers (depending on therapy administered) are associated with adverse outcome.(17, 18, 19, 20, 21)Also, reports suggest CD8 expression appears to be increased and macrophage density decreased in ESFL relative to ASFL.(10) A curated list of T-cell (*CD3D, CD4, CD8A, FOXP3*), macrophage (*CD68, CD163)* and macrophage-related (*CD47)* genes, were tested. Cut-point was determined by maximally selected rank statistics (applying a minimum group threshold of 15%) with p-values adjusted for multiple testing (Table S6).

Elevated intratumoural *CD8A* expression (n=26/97, 26.8%, Figure 4A) was associated with ∼2-fold increase in PFS for all patients HR=0.45, 95%CI=0.23-0.89, p=0.0042). Expression of the other immune genes were not associated with differential PFS. Importantly, elevated CD8A expression was confirmed as favourable in both the AusESFL (HR=0.45, 95%CI=0.16-0.93, p=0.038) and CanESFL (HR=0.35, 95%CI=0.10-1.14, p=0.038) validation cohorts (Figure 4B and C).

**Figure 4:**
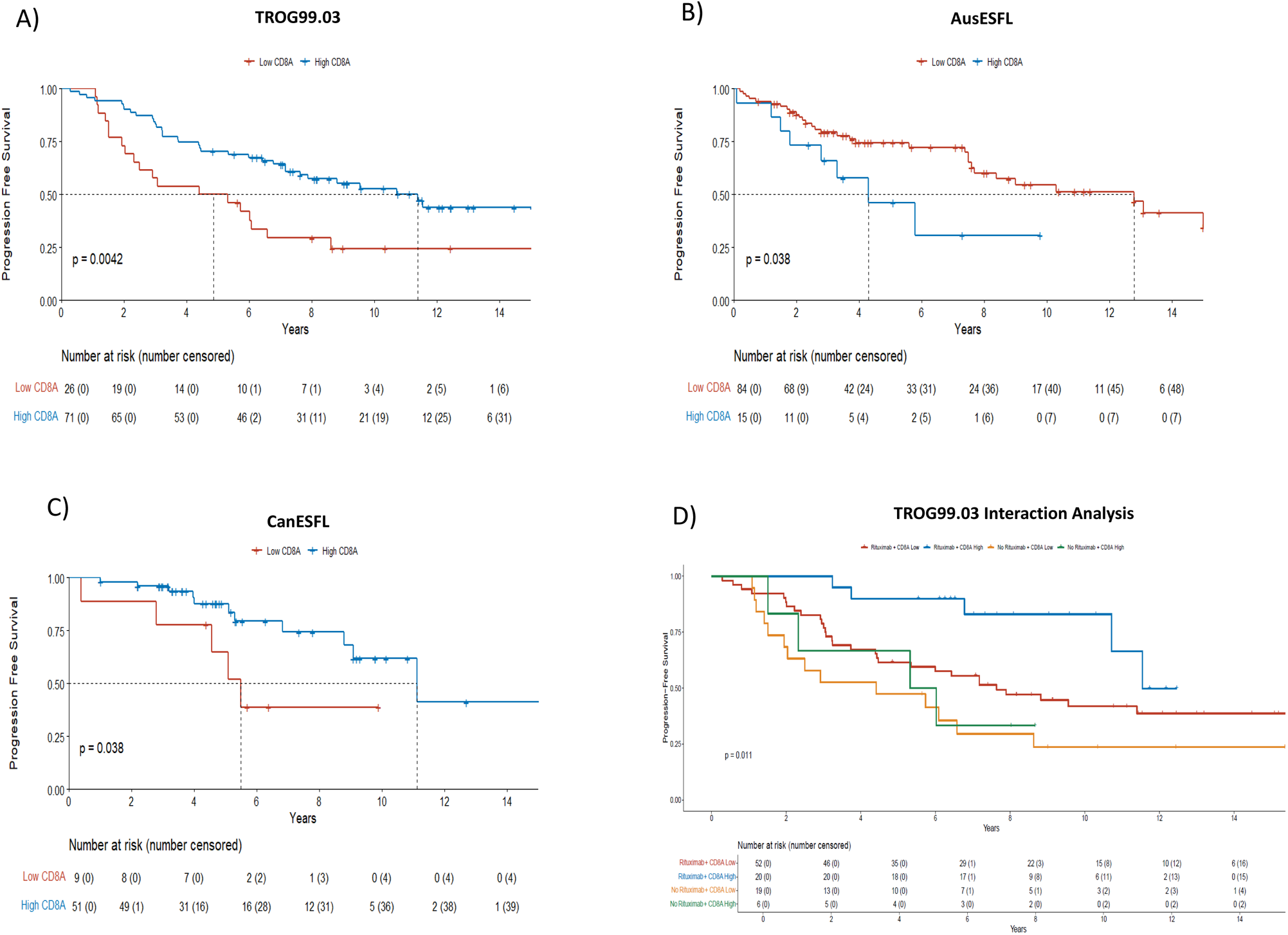
Progression-free survival stratified by CD8A gene expression in the discovery and validation cohorts, and CD8A / rituximab interaction analysis. A) TROG99.03 discovery cohort. B) AusESFL validation cohort. C) CanESFL validation cohort. D) TROG99. 03 discovery cohort stratified by rituximab use and CD8A gene expression.

There was no relationship between the presence of any genomic variants and CD8A gene expression (Figure S3). The impact of CD8A expression and of rituximab exposure on PFS was assessed by univariate analysis in the 97 patients. Both rituximab exposure and elevated CD8A were associated with improved PFS. Both rituximab exposure (HR=0.41, 95%CI=0.20-0.84, p=0.015) and elevated CD8A (HR=0.45, 95%CI=0.25-0.77, p=0.005) retained independent significance on multivariate analysis. We then performed Cox proportional hazards regression analysis to examine the interaction between rituximab and CD8A expression on PFS in the TROG99.03 cohort. However, the HR did not reach statistical significance (HR=0.54 95%CI=0.27-1.08, p=0.079). In contrast, by log-rank testing, the median PFS was significantly longer in those with high CD8A expression treated with rituximab (median 11.5yrs) than those without rituximab exposure (6.58yrs) and or those with rituximab exposure and low CD8A expression (5.67yrs) (Figure 4D). Put together, these data suggest but do not conclusively demonstrate a potential interaction between CD8A expression and rituximab.

### The density of CD8+ T-cells in intra-follicular areas relative to extra-follicular areas is higher in ESFL compared to ASFL

A curated panel of T-cell genes were compared between AusESFL and AusASFL (cohort description Table S7). Gene counts for *CD3D* (p=0.013), *CD4* (p=0.006) and *CD8A* (p=0.049) were increased in ESFL while markers of T follicular helper (*CXCR5*) and T regulatory (*FOXP3*) cells were similar to ASFL (Figure 5A).

**Figure 5:**
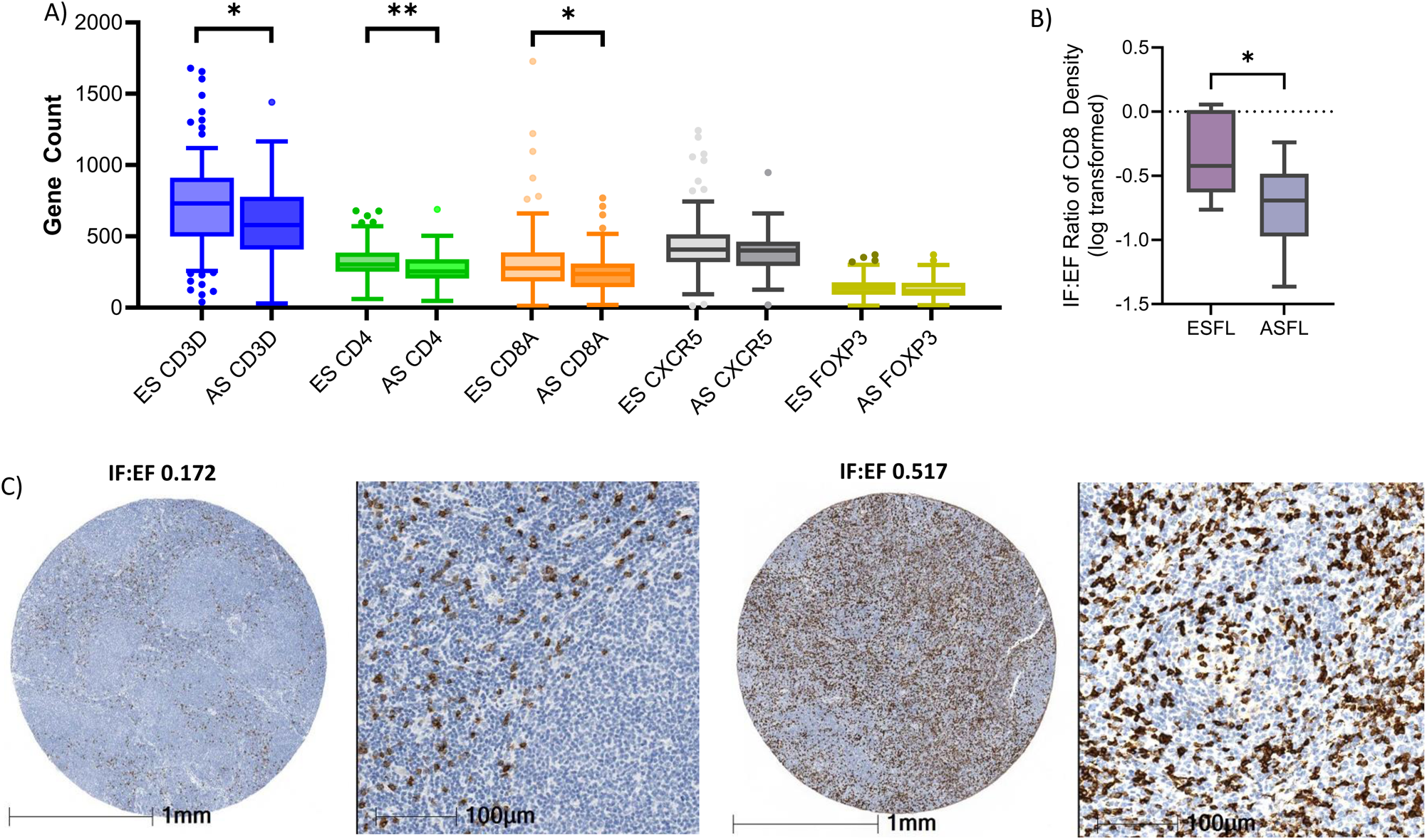
T-cell Enrichment in the ESFL tumour microenvironment. A) Digital gene expression of T-cell genes is increased in ESFL compared with ASFL. B) The CD8 intrafollicular:extrafollicular (IF:EF) ratio as measured by immunohistochemistry demonstrates significant enrichment by of CD8+ T-cells in interfollicular regions. C) Representative cases of low IF:EF ratio from diagnostic biopsy of an ASFL patient (left) and high IF:EF ratio from diagnostic biopsy of an ESFL (right) by light microscopy at 10x and 40x magnification.

Orthogonal validation was performed using immunohistochemistry on a subset of ESFL and ASFL with tissues available for TMA (21 ESFL, 53 ASFL). The expression density of CD8+ T-cells in intra-follicular areas relative to extra-follicular areas (IF:EF ratio, Figure 5B and C) was higher in ESFL than ASFL (p=0.047).

### HLA-class I specific neoantigens are present in ESFL

Next, we investigated the mechanistic relevance for the increased density of CD8+ T-cells in ESFL relative to ASFL, and why elevated CD8A transcriptomes levels might be associated with superior PFS. For this, the DNA sequencing data was re-interrogated to identify HLA-class I specific neoantigens. These are unique immunogenic self-antigens generated by tumour cells that arise because of the genomic mutations present within the malignant FL cell.(22) CD8+ T-cells recognise neoantigens in the context of HLA-class I molecules. We used an established high-stringency HLA-class I neoantigen-specific prediction pipeline,(14) which we validated using a combined approach incorporating confirmation of neoantigen peptide-expression, HLA-class I peptide binding assays, and γ-interferon secretion (supplemental).

The pipeline was applied to the 69 TROG99.03 diagnostic samples that had available germline DNA for HLA-class I genotyping (germline DNA was not available in the validation cohorts). Using the stringent criteria, 43% of samples had ≥1 HLA-class I neoantigens detected, with 55 neoantigens total. Neoantigens were found in a range of genes and included neoantigens derived from founder mutations such as *CREBBP* and *KMT2D.* Details of mutations leading to neoantigens are provided in Figure 6A. The majority of neoantigens were the result of missense mutations. In keeping with data derived in solid tumours, in ESFL we observed frameshift variants were most frequently observed for *KMT2D*.(23) Neoantigen data on 3 TROG99.03 patients with paired diagnostic/progressed samples are contained in the supplemental.

**Figure 6:**
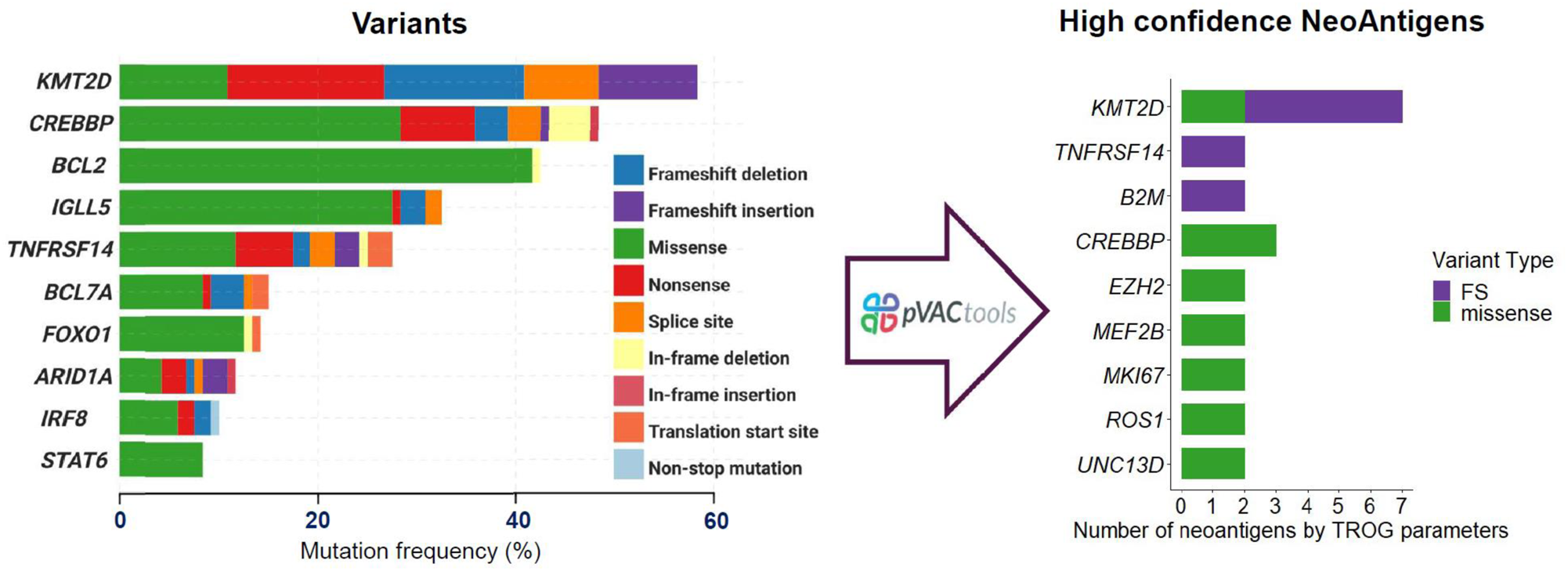
HLA-class I specific neoantigens in ESFL. Frequency of mutations in the TROG99.03 cases that had available germ-line DNA for HLA-class I genotyping, including ‘disruptive’ (i.e. nonsense and frameshift) and non-disruptive (predominantly missense) mutations. A histogram showing all genes with ≥2 neoantigens is shown in the right panel.

## Discussion

Herein we present extended follow-up of the TROG99.03 phase III RCT, including analyses of the impact of combining adjuvant rituximab with IFRT and chemotherapy on PFS. With a median follow-up of 11.3-years, treatment with rituximab-containing therapy (IFRT+R-CVP) was associated with greatly improved long-term PFS relative to non-rituximab regimens (IFRT alone or IFRT+CVP). Although no OS advantage was observed with systemic therapy, IFRT+(R)CVP treated patients experienced significantly fewer composite events (histological transformation and death). Independently of rituximab, elevated CD8A (validated in two independent PET-staged ESFL cohorts) was also associated with improved PFS. The AusESFL cohort were treated with a variety of different strategies, whereas all CanESFL patients had IFRT alone. Although this indicates the robustness of the findings, given the retrospective nature of the data collection no definitive conclusions regarding generalizability of the data can be made.

IFRT or involved-node RT (INRT) in the range 24-30Gy is associated with low toxicity and is currently preferred for curative-intent therapy.(24) Low-dose radiotherapy (4Gy) can usually attain local responses in indolent lymphoma but provides a lower probability of long-term local disease control than 24Gy.(25) Although ESFL is potentially curable with IFRT alone, relapses commonly occur outside irradiated volumes.(26) PET-staging improves accuracy of staging,(27) and is associated with better PFS.(26, 27, 28) Nevertheless, relapses still commonly occur outside irradiated volumes in PET-staged patients.

In the TROG99.03 study, PET-staged patients experienced the greatest benefit from adjuvant systemic therapy, indicating that systemic therapy improved outcomes by controlling occult systemic disease. The German phase II MIR study reported superior outcomes compared to historical series, when rituximab was given in addition to IFRT in ESFL.(29) Minimal residual disease analysis revealed a clonal marker in 36% of patients at diagnosis, with all but one marker positive patients attaining molecular remission after treatment.(29) In a retrospective real-world cohort, IFRT alone and systemic therapy without maintenance rituximab both yielded similar PFS.(8) However, in the same study, addition of maintenance rituximab improved PFS and was cost-effective.(8, 30). A randomised clinical trial recently commenced, comparing RT alone with RT plus rituximab (NCT01473628). More recently, the final analysis of the phase II Italian FIL MIRO study showed that in a uniformly PET-staged ESFL cohort, MRD-positive patients after IFRT were able to achieve MRD-negativity in 92% of cases with subsequent course of CD20 mAb ofatumumab.(6) In TROG99.03, adjuvant R-CVP given after IFRT was superior to non-rituximab regimens. The additional benefit of adding CVP chemotherapy to rituximab immunotherapy remains uncertain. The long duration of follow-up required for this trial in ESFL patients has led to potential confounding factors that cannot be entirely accounted for in the analysis of R-CVP treated patients as the healthcare landscape evolved over time. These include the introduction of FDG-PET as an important standard-of-care imaging modality and incremental gains in radiotherapy techniques leading to improved efficacy and long-term toxicity. A well-designed RCT comparing adjuvant rituximab after IFRT to IFRT plus rituximab plus chemotherapy in PET-staged ESFL would be required before definitively concluding that chemotherapy can safely be omitted. Nevertheless, adjuvant CD20 mAb alone, as per the MIR and FIL MIRO study, could be considered in addition to IFRT for ESFL patients who are unable to tolerate chemotherapy or are at increased risk of toxicity, and seek treatment with the highest probability of long-term MRD negativity and PFS.

Identification of risk factors in ESFL can assist with prognosis, management and future trial design. In TROG99.03, presence of extranodal disease (most likely explained by the inclusion of cases of duodenal FL)(15) approached significance for association with superior PFS. No other individual clinical factors, nor the FLIPI index, had prognostic utility. Neither *BCL2*-translocations, nor mutations in individual genes (including *BCL2, KMT2D* and *STAT6* mutations, were associated with PFS.

In ASFL, the importance of the TME is well-established.(16, 18, 31) The development of ASFL is dependent on the co-evolution of a tumour supportive niche,(32) while simultaneously FL B-cells must evade anti-tumour effector cells including CD8^+^ T-cells. This equilibrium is implicated in ASFL evolution, disease trajectory and treatment responses.(33, 34) Fittingly, most biomarker studies in FL rely on quantification of TME features rather than genomic alterations. In ASFL the TME has significant predictive impacts for the efficacy of rituximab where it abrogates the historically poor prognostic impact of pro-tumoural associated macrophages and confers favourable PFS in the setting of increased CD8 TIL infiltration.(16, 19, 35, 36, 37, 38, 39) This likely reflects rituximab-induced cell death and subsequent tumour antigen-specific immune response.(40) Recent studies have shown that intratumoural T-cell infiltration is influenced by interdependence of neoantigens, antigen-presentation and the broader TME composition.(18, 41, 42)

By contrast to ASFL, studies examining the TME of ESFL are sparse. Interestingly, the German Low Grade Study Group found differences in immune gene expression signatures (incorporating genes present to varying degrees across a range of lymphocyte lineages), between ESFL and ASFL.(12) Another recent study showed intratumoural CD8+ T-cells are more frequent within the TME of ESFL tissues versus ASFL.(10) In agreement with these published observations, the current study found CD8A transcriptomes were raised in ESFL compared to ASFL. Notably, intrafollicular CD8+ T-cell density was elevated in ESFL relative to ASFL.

Although genetic aberrations within the malignant FL cell were not directly associated with PFS, our data suggests they indirectly influence the composition of CD8+ T-cells within the TME via the production of neoantigens. These immunogenic peptides, when presented in the context of HLA-class I to intratumoural CD8+ T-cells, are known to be important in preventing systemic dissemination of malignancy and in mediating cancer control.(22) It has previously been shown that expanded populations of clonal CD8+ T-cells are present within the TME of FL, strongly suggestive of encounter with an antigenic stimulus.(21) Despite being a relatively low tumour mutation burden malignancy, HLA-class I neoantigens were detectable in 43% of paraffin-embedded tissues, including within driver mutations. It is likely that a higher detection rate would have been observed in fresh frozen tissues. Increased neoantigen-specific CD8+ T-cells may therefore explain the CD8 transcriptomic and IHC findings. Furthermore, neoantigens presented in the context of HLA-class I to intratumoural CD8+ T-cells may influence the favourable PFS in ESFL. In the small number of cases where relapsed/transformed ESFL samples were compared to their paired diagnostic tissue samples, neoantigens present at diagnosis remained detectable (and new neoantigens in one case were also detectable), indicating that immune-evasion had evolved to circumvent neoantigen-specific T-cell immunity. These data have implications not only for the pathogenesis of FL but also regarding the feasibility of neoantigen cancer vaccines, which remains an area of active research.

Comparisons of therapeutic approaches in ESFL are extremely difficult to conduct because of the relative rarity of localised disease and long natural history. In TROG99.03 the trial arms began to separate for PFS at around 5 years. Differences in overall survival were not statistically significant with a median follow up of over 11 years evidencing the significant duration of follow-up required to provide insights on improved curative potential or lymphoma-specific survival in this disease entity. The only prior RCT successfully completed in ESFL was the British National Lymphoma Investigation study. This showed no benefit from adding low-dose chlorambucil to RT.(43) Strengths of the current RCT are its extensive and near-complete follow-up, inclusion of PET-staging and rituximab from 2006Whilethe study is well powered for assessment of the primary endpoint, the relatively cohort remains relatively small for post-hoc and analysis and thus, it is difficult to make definitive conclusions around the predictive impacts of specific variables. Additionally, the prolonged accrual prevented evaluation of more potent anti-CD20 mAb, such as obinutuzumab and/or modern non-invasive risk-adapted approaches.(5, 44)

An additional strength of this study is the incorporation of translational studies on the prospectively accrued biospecimens, the application of clinically relevant techniques in formalin-fixed paraffin-embedded (FFPE) samples and access to independent validation cohorts. The long duration of the study is a challenge for biomarker collection and poses limitations on translational studies. Specifically, modern high-throughput techniques such as single-cell analysis require specific collection techniques which were not well described at the outset of the trial. Similarly, translational methods on biospecimens needed to be appropriately validated in samples collected over a long study duration to be applicable to the entire TROG99.03 cohort, as well as non-uniformly biospecimen collection in the validation cohorts. Prospective clinical trials incorporating such strategies that also include CD8 evaluation on the diagnostic biopsy would be invaluable in establishing robust, clinically-applicable biomarkers to guide treatment decisions. The TROG99.03 study indicates that collaborative group studies in ESFL, involving sustained clinical and radiographic follow-up, with centralized multi-omic analysis of biospecimens are feasible.

In summary, we present extended follow-up of the only phase III RCT of ESFL to include rituximab treated patients in which we demonstrate a significant improvement in PFS for those receiving adjuvant R-CVP after IFRT. Elevated intratumoural CD8 T-cells were independently associated with sustained disease control; future studies may establish a role for CD8 as a biomarker to guide risk-stratified application of adjuvant therapy after IFRT. CD8+ T-cell neoantigens were frequently detectable suggesting neoantigen-specific CD8+ T-cells have a role in confining the spread of the disease. The development of novel neoantigen-directed treatments for ESFL may leverage these findings to produce patient-specific therapies in the future. These data have important implications for the current management of ESFL and the design of prospective clinical trials.

## Data Availability

produced in the present study are available upon reasonable request to the authors

## Acknowledgements

This study was supported by Cancer Council Victoria, the Australasian Leukaemia Lymphoma Group, National Health and Medical Research Council and the Translational Molecular Pathology-Immunoprofiling (TMP-IL) Moonshots Platform Halo AI capabilities at the Department Translational Molecular Pathology, the University of Texas MD Anderson Cancer Center. Slide scanning and laboratory work: Luisa Solis, Mei Jiang, Beatriz Sanchez-Espiridion, Wei Lu, Khaja Khan, and Jianling Zhou.

## Contributors

MPM, JPS, JTWD, and MKG contributed to study conception and design. MPM, JPS, DR, POB, AM, RT, SD, and DC were principal investigators of the clinical trial and contributed to study initiation. HT, CK, PB, ERT, JG, SCL, MBS, KN, SHK, MMP, DED, and MRG performed experiments and/or interpreted data. RK, VS, SH, TB, DL, NJ, MB, ML, FdA, CC, BA, TC, JB, AJ, MS, LL, and CV enabled access to validation cohorts and curated data. The manuscript was written and compiled by MPM, JTWD, and MKG, who also accessed and verified the data. All authors contributed to revision and finalisation of the paper, and the decision to submit for publication.

## Data sharing

Patient data will be provided by TROG on request. Any request should be sent to MMM, with a detailed description of the research protocol. TROG reserves the right to decide whether to share the data or not. Access will be provided after a proposal has been approved by an independent review committee established for this purpose and after receipt of a signed data sharing agreement.

## Conflict of Interest Statement

Amgen (GCSF) and Roche (Rituximab) for the provision of study materials. There are no other relevant conflicts of interest.

## Notes

### Competing Interest Statement

The authors have declared no competing interest.

### Clinical Trial

NCT00115700

### Clinical Protocols

https://doi.org/10.1200/JCO.2018.77.9892

### Funding Statement

This trial received primary funding from the Cancer Council of Victoria with additional funding contributed by the Australasian Leukaemia and Lymphoma Group Peter MacCallum Cancer Centre Amgen and Roche. Roche also supplied the rituximab used in the amended version of the trial. QA for the trial was supported by a grant from the Australian Federal Department of Health.

### Author Declarations

Human research ethics committee of Peter MacCallum Cancer Centre have ethical approval for this work

## References

1. Teras LR, DeSantis CE, Cerhan JR, Morton LM, Jemal A, Flowers CR. 2016 US lymphoid malignancy statistics by World Health Organization subtypes. CA Cancer J Clin. 2016;66(6):443–59.

2. Smith A, Crouch S, Lax S, Li J, Painter D, Howell D, et al. Lymphoma incidence, survival and prevalence 2004-2014: sub-type analyses from the UK’s Haematological Malignancy Research Network. Br J Cancer. 2015;112(9):1575–84.

3. van Leeuwen MT, Turner JJ, Joske DJ, Falster MO, Srasuebkul P, Meagher NS, et al. Lymphoid neoplasm incidence by WHO subtype in Australia 1982-2006. Int J Cancer. 2014;135(9):2146–56.

4. Wirth A, Mikhaeel NG, Aleman BMP, Pinnix CC, Constine LS, Ricardi U, et al. Involved Site Radiation Therapy in Adult Lymphomas: An Overview of International Lymphoma Radiation Oncology Group Guidelines. Int J Radiat Oncol Biol Phys. 2020;107(5):909–33.

5. Marcus R, Davies A, Ando K, Klapper W, Opat S, Owen C, et al. Obinutuzumab for the First-Line Treatment of Follicular Lymphoma. N Engl J Med. 2017;377(14):1331–44.

6. Pulsoni A, Ferrero S, Tosti ME, Luminari S, Dondi A, Cavallo F, et al. Local radiotherapy and measurable residual disease-driven immunotherapy in patients with early-stage follicular lymphoma (FIL MIRO): final results of a prospective, multicentre, phase 2 trial. Lancet Haematol. 2024;11(7):e499–e509.

7. MacManus MP, Hoppe RT. Is radiotherapy curative for stage I and II low-grade follicular lymphoma? Results of a long-term follow-up study of patients treated at Stanford University. J Clin Oncol. 1996;14(4):1282–90.

8. Tobin JWD, Rule G, Colvin K, Calvente L, Hodgson D, Bell S, et al. Outcomes of stage I/II follicular lymphoma in the PET era: an international study from the Australian Lymphoma Alliance. Blood Adv. 2019;3(19):2804–11.

9. MacManus M, Fisher R, Roos D, O’Brien P, Macann A, Davis S, et al. Randomized Trial of Systemic Therapy After Involved-Field Radiotherapy in Patients With Early-Stage Follicular Lymphoma: TROG 99.03. J Clin Oncol. 2018:JCO2018779892.

10. Los-de Vries GT, Stevens WBC, van Dijk EV, Langois-Jacques C, Clear AJ, Stathi P, et al. Genomic and microenvironmental landscape of stage I follicular lymphoma, compared to stage III/IV. Blood Adv. 2022.

11. Kalmbach S, Grau M, Zapukhlyak M, Leich E, Jurinovic V, Hoster E, et al. Novel insights into the pathogenesis of follicular lymphoma by molecular profiling of localized and systemic disease forms. Leukemia. 2023;37(10):2058–65.

12. Staiger AM, Hoster E, Jurinovic V, Winter S, Leich E, Kalla C, et al. Localized- and advanced-stage follicular lymphomas differ in their gene expression profiles. Blood. 2020;135(3):181–90.

13. Nath K, Law SC, Sabdia MB, Gunawardana J, de Long LM, Sester D, et al. Intratumoral T cells have a differential impact on FDG-PET parameters in follicular lymphoma. Blood Adv. 2021;5(12):2644–9.

14. Wells DK, van Buuren MM, Dang KK, Hubbard-Lucey VM, Sheehan KCF, Campbell KM, et al. Key Parameters of Tumor Epitope Immunogenicity Revealed Through a Consortium Approach Improve Neoantigen Prediction. Cell. 2020;183(3):818–34 e13.

15. Hellmuth JC, Louissaint A, Jr., Szczepanowski M, Haebe S, Pastore A, Alig S, et al. Duodenal-type and nodal follicular lymphomas differ by their immune microenvironment rather than their mutation profiles. Blood. 2018;132(16):1695–702.

16. Stevens WBC, Mendeville M, Redd R, Clear AJ, Bladergroen R, Calaminici M, et al. Prognostic relevance of CD163 and CD8 combined with EZH2 and gain of chromosome 18 in follicular lymphoma: a study by the Lunenburg Lymphoma Biomarker Consortium. Haematologica. 2017;102(8):1413–23.

17. Bolen CR, McCord R, Huet S, Frampton GM, Bourgon R, Jardin F, et al. Mutation load and an effector T-cell gene signature may distinguish immunologically distinct and clinically relevant lymphoma subsets. Blood Adv. 2017;1(22):1884–90.

18. Han G, Deng Q, Marques-Piubelli ML, Dai E, Dang M, Ma MCJ, et al. Follicular Lymphoma Microenvironment Characteristics Associated with Tumor Cell Mutations and MHC Class II Expression. Blood Cancer Discov. 2022;3(5):428–43.

19. Kridel R, Xerri L, Gelas-Dore B, Tan K, Feugier P, Vawda A, et al. The Prognostic Impact of CD163-Positive Macrophages in Follicular Lymphoma: A Study from the BC Cancer Agency and the Lymphoma Study Association. Clin Cancer Res. 2015;21(15):3428–35.

20. Lee AM, Clear AJ, Calaminici M, Davies AJ, Jordan S, MacDougall F, et al. Number of CD4+ cells and location of forkhead box protein P3-positive cells in diagnostic follicular lymphoma tissue microarrays correlates with outcome. J Clin Oncol. 2006;24(31):5052–9.

21. Tobin JWD, Keane C, Gunawardana J, Mollee P, Birch S, Hoang T, et al. Progression of Disease Within 24 Months in Follicular Lymphoma Is Associated With Reduced Intratumoral Immune Infiltration. J Clin Oncol. 2019:JCO1802365.

22. Sabdia MB, Patch AM, Tsang H, Gandhi MK. Neoantigens - the next frontier in precision immunotherapy for B-cell lymphoproliferative disorders. Blood Rev. 2022;56:100969.

23. Koster J, Plasterk RHA. A library of Neo Open Reading Frame peptides (NOPs) as a sustainable resource of common neoantigens in up to 50% of cancer patients. Sci Rep. 2019;9(1):6577.

24. Campbell BA, Voss N, Woods R, Gascoyne RD, Morris J, Pickles T, et al. Long-term outcomes for patients with limited stage follicular lymphoma: involved regional radiotherapy versus involved node radiotherapy. Cancer. 2010;116(16):3797–806.

25. Lowry L, Smith P, Qian W, Falk S, Benstead K, Illidge T, et al. Reduced dose radiotherapy for local control in non-Hodgkin lymphoma: a randomised phase III trial. Radiother Oncol. 2011;100(1):86–92.

26. Hoskin PJ, Kirkwood AA, Popova B, Smith P, Robinson M, Gallop-Evans E, et al. 4 Gy versus 24 Gy radiotherapy for patients with indolent lymphoma (FORT): a randomised phase 3 non-inferiority trial. Lancet Oncol. 2014;15(4):457–63.

27. Wirth A, Foo M, Seymour JF, Macmanus MP, Hicks RJ. Impact of [18f] fluorodeoxyglucose positron emission tomography on staging and management of early-stage follicular non-hodgkin lymphoma. Int J Radiat Oncol Biol Phys. 2008;71(1):213–9.

28. Brady JL, Binkley MS, Hajj C, Chelius M, Chau K, Balogh A, et al. Definitive radiotherapy for localized follicular lymphoma staged by (18)F-FDG PET-CT: a collaborative study by ILROG. Blood. 2019;133(3):237–45.

29. Herfarth K, Borchmann P, Schnaidt S, Hohloch K, Budach V, Engelhard M, et al. Rituximab With Involved Field Irradiation for Early-stage Nodal Follicular Lymphoma: Results of the MIR Study. Hemasphere. 2018;2(6):e160.

30. Tobin JWD, Crothers A, Ma TE, Mollee P, Gandhi MK, Scuffham P, et al. A cost-effectiveness analysis of front-line treatment strategies in early-stage follicular lymphoma. Leuk Lymphoma. 2021:1–9.

31. Huet S, Tesson B, Jais JP, Feldman AL, Magnano L, Thomas E, et al. A gene-expression profiling score for prediction of outcome in patients with follicular lymphoma: a retrospective training and validation analysis in three international cohorts. Lancet Oncol. 2018;19(4):549–61.

32. Ame-Thomas P, Maby-El Hajjami H, Monvoisin C, Jean R, Monnier D, Caulet-Maugendre S, et al. Human mesenchymal stem cells isolated from bone marrow and lymphoid organs support tumor B-cell growth: role of stromal cells in follicular lymphoma pathogenesis. Blood. 2007;109(2):693–702.

33. Tobin JWD, Gandhi MK. Discordant solutions to discordant problems. Blood. 2021;137(21):2857–8.

34. Haebe S, Shree T, Sathe A, Day G, Czerwinski DK, Grimes SM, et al. Single-cell analysis can define distinct evolution of tumor sites in follicular lymphoma. Blood. 2021;137(21):2869–80.

35. de Jong D, Koster A, Hagenbeek A, Raemaekers J, Veldhuizen D, Heisterkamp S, et al. Impact of the tumor microenvironment on prognosis in follicular lymphoma is dependent on specific treatment protocols. Haematologica. 2009;94(1):70–7.

36. Takahashi H, Tomita N, Sakata S, Tsuyama N, Hashimoto C, Ohshima R, et al. Prognostic significance of programmed cell death-1-positive cells in follicular lymphoma patients may alter in the rituximab era. Eur J Haematol. 2013;90(4):286–90.

37. Laurent C, Flores M, Chartier L, Huet S, Bolen CR, Venstrom JM, et al. Long-term follow-up confirms the favourable prognostic impact of high numbers of tumour infiltrating CD3 T-cells in follicular lymphoma patients treated by rituximab-maintenance regimen. Br J Haematol. 2023;202(3):686–9.

38. Laurent C, Muller S, Do C, Al-Saati T, Allart S, Larocca LM, et al. Distribution, function, and prognostic value of cytotoxic T lymphocytes in follicular lymphoma: a 3-D tissue-imaging study. Blood. 2011;118(20):5371–9.

39. Alvaro T, Lejeune M, Salvado MT, Lopez C, Jaen J, Bosch R, et al. Immunohistochemical patterns of reactive microenvironment are associated with clinicobiologic behavior in follicular lymphoma patients. J Clin Oncol. 2006;24(34):5350–7.

40. Cheadle EJ, Sidon L, Dovedi SJ, Melis MH, Alduaij W, Illidge TM, et al. The induction of immunogenic cell death by type II anti-CD20 monoclonal antibodies has mechanistic differences compared with type I rituximab. Br J Haematol. 2013;162(6):842–5.

41. Mondello P, Fama A, Larson MC, Feldman AL, Villasboas JC, Yang ZZ, et al. Lack of intrafollicular memory CD4 + T cells is predictive of early clinical failure in newly diagnosed follicular lymphoma. Blood Cancer J. 2021;11(7):130.

42. Radtke AJ, Postovalova E, Varlamova A, Bagaev A, Sorokina M, Kudryashova O, et al. Multi-omic profiling of follicular lymphoma reveals changes in tissue architecture and enhanced stromal remodeling in high-risk patients. Cancer Cell. 2024;42(3):444–63 e10.

43. Kelsey SM, Newland AC, Hudson GV, Jelliffe AM. A British National Lymphoma Investigation randomised trial of single agent chlorambucil plus radiotherapy versus radiotherapy alone in low grade, localised non-Hodgkins lymphoma. Med Oncol. 1994;11(1):19–25.

44. Schroers-Martin JG, Alig S, Garofalo A, Tessoulin B, Sugio T, Alizadeh AA. Molecular Monitoring of Lymphomas. Annu Rev Pathol. 2023;18:149–80.

