## Supplemental Data & Tables for "Adjuvant rituximab and elevated intratumoural CD8 expression are associated with sustained disease control after radiotherapy in early-stage follicular lymphoma: TROG99.03"

**Index**


| | |
|---|---|
| **P2** | **Supplemental Methods** |
| **P3** | **Supplemental Results** |
| **P4** | **Supplemental References** |
| **P5** | **Supplemental Tables** |
| **P12** | **Supplemental Figures** |

**Supplemental Methods**

**Data Management & Confidentiality**

Trial data and confidentiality measures are outlined in the study protocol. De-identified data may potentially be shared for secondary analyses upon a formal written request to TROG. Such analyses would require a detailed protocol with appropriate institutional review board approval and review and approval by the TROG secondary analysis committee.

**Gene expression and DNA profiling**

The NanoString® 770-gene PanCancer Immune panel (730 immune-related genes plus 40 housekeeping genes) was used to digitally quantify genes as previously published.(1) Gene expression data was normalised via the geNorm algorithm incorporated into the nSolver 4.0 software. For sequencing a minimum of 200ng of DNA was used, with a 330 gene targeted 'PanHaem V2.0' panel.(2) Variant annotation and filtering were performed with PathOS.(3) Variants detected in >35% of all samples tested on PanHaem V2.0 panel were excluded. These common variants were assumed to be either artefacts or polymorphisms. Variants present in more than 1% of the population, as identified by the 1000 Genomes Project, were excluded. Variants as determined by the Ensembl Variant Effect Predictor, were excluded.(4) Variants that were either established pathogenic variants, detected at a somatic variant allele frequency, not reported in gnomAD, are not panel artefacts and all BCL2 variants were retained. Filtered Variant Call Files (VCFs) were converted to Mutation Annotation Format (MAFs) using vcf2maf tool (v1.6.21). Variant analysis was performed on MAF files with the R package 'MAFtools'.(5) BCL2-translocation detection was performed with GRIDDS (v2.2.3).(6) For the FL samples used to functionally validate neoantigen methodology, a custom hybridisation capture panel designed by the Australian Translational Genomics Centre panel was used.

**Immunohistochemistry**

Sections (4μm) were prepared from paraffin-embedded tissue microarray (TMA) samples with 1mm diameter/cores. Slides were stained using Leica Bond RX slide stainer (Leica Biosystems) following an optimized protocol per antibody. For single T-cell marker detection, antigen retrieval used Bond ER Solution (Leica Biosystems) for 60 minutes at $21^{0}$C. CD8 primary antibody (Dako M7103) diluted 1:100 was detected using Bond Polymer Refine Detection kit with diaminobenzidine as chromogen. Slides were counterstained with haematoxylin. Tonsil and appendix tissues sections were used as positive controls.
Automated digital image analysis used an Aperio AT2 slide scanner (Leica Biosystems) at 20x magnification, and a pathologist (blinded to clinical data) segmented the tissue into intra-follicular and extra-follicular areas. The membrane/cytoplasmic expression of CD8 was evaluated and scored according to the intensity of expression (0, negative; 1+, weak; 2+, moderate; and 3+, strong). Cell density was obtained using HALO v2.3 image analysis software (Indica Labs) using the Cytonuclear v1.6 algorithm and an H-score calculated.

**Neoantigen profiling**

4-digit HLA-class I genotyping was performed on germline (blood) samples using Optitype and HLA-HD,(7, 8) (with discordant results omitted from the analysis).
Filtered VCFs as per above, were used for neoantigen calling. Neoantigens were predicted with pVACseq (v1.5.9).(9) All options remained as default apart from use of all 8 HLA class-I prediction algorithms, binding threshold of 50nM, 'Best MT Score' binding filter, and mutant (MT) peptide binding affinity less than wild type (WT). Additionally, neoantigens were further filtered based on 'Tumor Neoantigen Selection Alliance' (TESLA) consortium guidelines,(10) to include candidates with agreptopicity>0.1 (MT binding affinity/WT binding affinity >0.1). Neoantigen candidates were aligned to using the Protein Basic Local Alignment Search Tool (BLAST), to identify if neoantigens were present in the normal proteome.(4, 9, 10) Functional validation of neoantigen calling is in supplemental results.

**RT-PCR and Sanger sequencing**

To confirm RNA expression of mutations creating neoantigens, reverse transcription polymerase chain reaction (RT-PCR) and Sanger sequencing were used. Briefly, cDNA was synthesized from RNA obtained from FL tumour biopsies using SuperScript IV (Invitrogen). cDNA was amplified using Phusion Hi-Fidelity DNA Polymerase (Thermo Scientific) and primers flanking the mutation of interest, with primers spanning different exons to ensure amplification of cDNA transcripts. Amplified cDNA was gel purified and Sanger sequencing was performed by the Australian Genome Research Facility.

### Supplemental Results

#### Protocol Deviations

Major deviations from protocol treatment were as follows: One arm A patient was found to have more extensive disease after randomisation and received R-CVP but not IFRT. One arm A patient received wide-field radiation for more extensive disease. Two arm B patients randomised to CVP actually received R-CVP. One arm B patient had no trial treatment and pursued alternative medicine but was included in follow-up. Two arm B patients refused chemotherapy, one did not receive chemotherapy due to rheumatic fever, one had no chemotherapy because of interstitial lung disease and one arm B patient required hernia surgery and received no chemotherapy due to unhealed wound. One arm B patient received chemotherapy before IFRT.

#### Genomic and transcriptomic results

Alternative translocation partners for IgH locus were identified in 7 cases, including *BCL6,* and two instances of t14;16 translocation partnering with *CIITA* (a regulator of the HLA-class II complex) which to our knowledge has not previously been described.

To determine the relationship between genomic variants and CD8A gene expression, in the discovery cohort, forest plots of variants from 9 genes of interest (*KMT2D*, *BCL2* and the M7-FLIPI genes: *EZH2, ARID1A, MEF2B, EP300, FOXO1, CREBBP* and *CARD11*) were plotted according to maximally selected CD8A expression status (high, low). Adjusting for multiple variables, CD8A was independent of all mutations (Figure S3).

Following the initial mutational and nanoString analysis, only three TROG99.03 discovery cases had remaining tissue to confirm neoantigen RNA expression by RT-PCR, with Sanger sequencing used to distinguish mutant neoantigen (MT) from wild-type RNA expression. Notably, one of these had both diagnostic and relapsed samples. In that case, a neoantigen caused by a missense mutation in P2RY8 was predicted in the diagnostic sample. RT-PCR confirmed the presence of MT RNA in both samples, indicating that the neoantigen was expressed at both diagnosis and relapse. In the other two cases (with only a diagnostic sample available), neoantigens caused by frameshift mutations in TNFRSF14 (case 2) and KMT2D (case 3) were predicted. Expression of both MT RNA were individually confirmed by RT-PCR and Sanger sequencing.

#### Validation of *in-silico* neoantigen prediction methodology

As the TROG99.03 discovery cohort did not have peripheral blood mononuclear cells (PBMC) samples available, functional validation of the pipeline was performed in two samples obtained from the Princess Alexandra Hospital, Brisbane, in whom paired FL tissue and cryopreserved pre-therapy PBMC were available Firstly, HLA-lass I neoantigens were predicted (as per TROG9.03 samples) using the pVACtools pipeline. Then, a combined approach was adopted: i) RNA was extracted from FL tumour biopsies to confirm that neoantigen expression could be confirmed by RT-PCR and Sanger sequencing; ii) To confirm that neoantigens were capable of binding to HLA-class I molecules, an HLA-class I peptide binding assay was used (ProImmune REVEAL, ProImmune Ltd., Oxford, United Kingdom); iii) Binding to HLA-class I molecules was compared to that of a known relevant T-cell epitope, with very strong binding properties and a binding score for each HLA-class I-peptide complex calculated by comparison to the positive control (binding scores of >45 were considered positive); iv) Enzyme-linked immunospot (ELISPOT) assay was used (fold-change over negative control ≥1.5 defined as positive).

In both cases, RT-PCR and Sanger sequencing expression of this MT RNA confirmed that the predicted FL neoantigens (an insertion causing a frame-shift mutation in *NT5C2* and a missense mutation in *SETD8* respectively) were expressed within the tumours. In each, the REVEAL scores were >45 (69.2 and 49.8 respectively) indicating that the predicted FL neoantigen peptides were capable of binding to relevant HLA-lass I alleles for presentation to CD8+ T-cells. By ELISPOT, the predicted FL neoantigen peptides showed fold-change over negative control of 1.7 and 2.9 respectively, indicating the peptides were functionally able to induce interferon-g CD8+ T-cell release.

#### Neoantigens in disease progression

Finally, samples were available in 3 pairs of diagnostic vs. progressed ESFL tissues (2 relapsed, 1 transformed). In 1 patient no neoantigens were detected at diagnosis or relapse. In the other relapsed patient, a neoantigen was detected at diagnosis in *KMT2D* that was retained at relapse, along with the new detection of neoantigens in *FOXO1* and *TP53*. In the patient that transformed, 4 neoantigens were present at diagnosis (2 in *MEF2B*, 1 in *CRLF2* and 1 in *P2RY8*), with all 4 neoantigens remaining detectable in the transformed tissue.

**Supplemental References**

1. Nath K, Law SC, Sabdia MB, Gunawardana J, de Long LM, Sester D, et al. Intratumoral T cells have a differential impact on FDG-PET parameters in follicular lymphoma. Blood Adv. 2021;5(12):2644-9.
2. Blombery PA, Ryland GL, Markham J, Guinto J, Wall M, McBean M, et al. Detection of clinically relevant early genomic lesions in B-cell malignancies from circulating tumour DNA using a single hybridisation-based next generation sequencing assay. Br J Haematol. 2017.
3. Doig KD, Fellowes A, Bell AH, Seleznev A, Ma D, Ellul J, et al. PathOS: a decision support system for reporting high throughput sequencing of cancers in clinical diagnostic laboratories. Genome Med. 2017;9(1):38.
4. McLaren W, Gil L, Hunt SE, Riat HS, Ritchie GR, Thormann A, et al. The Ensembl Variant Effect Predictor. Genome Biol. 2016;17(1):122.
5. Mayakonda A, Lin DC, Assenov Y, Plass C, Koeffler HP. Maftools: efficient and comprehensive analysis of somatic variants in cancer. Genome Res. 2018;28(11):1747-56.
6. Cameron DL, Schroder J, Penington JS, Do H, Molania R, Dobrovic A, et al. GRIDSS: sensitive and specific genomic rearrangement detection using positional de Bruijn graph assembly. Genome Res. 2017;27(12):2050-60.
7. Szolek A, Schubert B, Mohr C, Sturm M, Feldhahn M, Kohlbacher O. OptiType: precision HLA typing from next-generation sequencing data. Bioinformatics. 2014;30(23):3310-6.
8. Kawaguchi S, Higasa K, Shimizu M, Yamada R, Matsuda F. HLA-HD: An accurate HLA typing algorithm for next-generation sequencing data. Hum Mutat. 2017;38(7):788-97.
9. Hundal J, Kiwala S, McMichael J, Miller CA, Xia H, Wollam AT, et al. pVACtools: A Computational Toolkit to Identify and Visualize Cancer Neoantigens. Cancer Immunol Res. 2020;8(3):409-20.
10. Wells DK, van Buuren MM, Dang KK, Hubbard-Lucey VM, Sheehan KCF, Campbell KM, et al. Key Parameters of Tumor Epitope Immunogenicity Revealed Through a Consortium Approach Improve Neoantigen Prediction. Cell. 2020;183(3):818-34 e13.

**Table S1: Clinical characteristics of TROG99.03 Clinical Cohort.**
Abbreviations: LDH, lactate dehydrogenase; B2M, beta-2-macroglobulin; ULN, upper limit normal.

| Patient Characteristic | | All TROG99.03 | | CMT | | IFRT | |
|---|---|---|---|---|---|---|---|
| | | N | % | N | % | N | % |
| All Patients | | 150 | - | 75 | - | 75 | - |
| Study period | Prior to 2006 Amendment | 88/150 | 59% | 44/75 | 59% | 44/75 | 59% |
| Sex | Male | 78/150 | 52% | 40/75 | 53% | 38/75 | 51% |
| Age | $\geq$60 | 68/150 | 45% | 34/75 | 45% | 34/75 | 45% |
| Stage | 1 | 113/150 | 75% | 56/75 | 75% | 57/75 | 76% |
| FDG-PET Staging | Yes | 72/150 | 48% | 35/75 | 47% | 37/75 | 49% |
| Extranodal Site | Yes | 12/150 | 8% | 7/75 | 9% | 5/75 | 7% |
| Histologic Grade | 1-2 | 143/147 | 97% | 73/74 | 99% | 70/73 | 96% |
| LDH | Raised > ULN | 16/143 | 11% | 5/71 | 7% | 11/72 | 15% |
| B2M | Raised > ULN | 12/124 | 10% | 5/63 | 8% | 7/61 | 11% |
| Bulky Disease | >5cm | 21/150 | 14% | 10/75 | 13% | 11/75 | 15% |

**Table S2: Clinical characteristics of Discovery and Validation cohorts**

AusESFL combined modality treatment (CMT) backbones included CHOP, CVP, bendamustine, chlorambucil and rituximab monotherapy. CD20 mAb in the AusESFL cohort included rituximab (40/44) and obinutuzumab (4/44). ‘Watchful Waiting’ patients who were not treated within 12 months of diagnosis were excluded from PFS analysis.

| Patient Characteristics | | All TROG99.03 | | TROG Discovery | | AusESFL Validation | | CanESFL Validation | |
|---|---|---|---|---|---|---|---|---|---|
| | | N | % | N | % | N | % | N | % |
| All Patients | | 150 | - | 101 | - | 99 | - | 72 | - |
| Study period | Prior to 2006 Amendment | 88/150 | 59 | 48/101 | 48 | N/A | - | NA | - |
| Sex | Male | 78/150 | 52 | 52/101 | 51 | 50/99 | 51 | NA | - |
| Age | ≥60 | 68/150 | 45 | 47/101 | 47 | 56/99 | 56 | 36/72 | 50 |
| Stage | 1 | 113/150 | 75 | 78/101 | 77 | 60/99 | 61 | 67/72 | 86 |
| FDG-PET Staging | Yes | 72/150 | 48 | 51/101 | 51 | 99/99 | 100 | 72/72 | 100 |
| Extranodal Site | Yes | 12/150 | 8 | 5/101 | 5 | 21/99 | 21 | 15/72 | 21 |
| Histologic Grade | 1-2 | 143/147 | 97 | 94/99 | 95 | 90/99 | 91 | NA | - |
| LDH | Raised > ULN | 16/143 | 11 | 13/93 | 13 | 12/99 | 12 | 9/67 | 13 |
| B2M | Raised > ULN | 12/124 | 10 | 6/81 | 93 | 11/63 | 17 | NA | - |
| Treatment | Watchful waiting | 0/150 | 0 | 0/101 | 0 | 15/99 | 15 | 0 | 0 |
| | Radiotherapy Alone | 75/150 | 50 | 49/101 | 49 | 36/99 | 36 | 72/72 | 100 |
| | CMT without CD20 mAb | 43/150 | 29 | 24/101 | 24 | 0/99 | 0 | 0/72 | 0 |
| | CMT with CD20 mAb | 32/150 | 21 | 28/101 | 28 | 48/99 | 49 | 0/72 | 0 |

**Table S3: Rates of malignancy in CMT vs IFRT arms**

| IFRT Arm | Combined Modality Treatment Arm |
|---|---|
| Skin Cancers n = 4<br>Melanoma = 2<br>Non Melanoma Skin Cancers = 2 | Skin Cancer n = 6<br>Melanoma = 0<br>Non Melanoma Skin Cancers = 6 |
| Solid Cancers n = 9<br>Prostate n = 2<br>Gastric/Oesophageal = 2<br>NSMCL = 2<br>Breast = 1<br>Bowel = 1<br>Other = 1 | Solid Cancer n = 9<br>Prostate n = 3<br>Gastric/Oesophageal = 1<br>NSMCL = 2<br>Breast = 1<br>Bowel = 1<br>Other = 1 |
| Haematological Cancer n = 0 | Haematological Cancer n = 1<br>Myelodysplasia = 1 |

**Table S4: Univariate and Multivariate Clinical Prognosticators for progression free survival in TROG99.03 study**

| | Univariate | | | Multivariate | | |
|---|---|---|---|---|---|---|
| | **p.value** | **HR** | **95% CI** | **p.value** | **HR** | **95% CI** |
| **Male** | 0.960 | 0.99 | (0.62-1.6) | 0.846 | 1.06 | (0.61-1.85) |
| **Stage II** | 0.200 | 1.40 | (0.84-2.3) | 0.478 | 1.25 | (0.67-2.34) |
| **Grade 3a** | 0.130 | 1.00 | (0.99-1) | 0.546 | 1.00 | (0.99-1.01) |
| **Age>60** | 0.750 | 0.93 | (0.58-1.5) | 0.769 | 1.09 | (0.61-1.97) |
| **LDH Elevated** | 0.970 | 0.98 | (0.45-2.2) | 0.709 | 1.18 | (0.49-2.79) |
| **B2M Elevated** | 0.046 | 0.47 | (0.22-0.99) | 0.146 | 0.53 | (0.23-1.25) |
| **Extranodal** | 0.041 | 0.13 | (0.018-0.91) | 0.050 | 0.14 | (0.02-1.01) |

**Table S5: Association between FL-related mutations and PFS and OS**

10 genes were mutated in ≥10% cases (frequencies are provided in Figure 3). The tables below show all 10 genes (ranked by p values) tested for association with PFS and OS calculated by univariable Cox regression. There was no significant association with PFS or OS after correcting for multiple testing (p values shown reflect un-corrected values).

| | PFS | | |
|---|---|---|---|
| Gene | P value | HR | 95% CI |
| TNFRSF14 | 0.085 | 0.56 | 0.91-3.47 |
| KMT2D | 0.157 | 1.48 | 0.39-1.17 |
| IRF8 | 0.264 | 0.56 | 0.64-4.92 |
| STAT6 | 0.317 | 1.47 | 0.32-1.45 |
| CREBBP | 0.352 | 0.77 | 0.75-2.25 |
| BCL7A | 0.384 | 1.36 | 0.37-1.47 |
| ARID1A | 0.713 | 0.84 | 0.47-2.99 |
| FOXO1 | 0.732 | 1.15 | 0.39-1.95 |
| IGLL5 | 0.753 | 1.11 | 0.47-1.72 |
| BCL2 | 0.831 | 0.94 | 0.59-1.92 |

| | OS | | |
|---|---|---|---|
| Gene | P value | HR | 95% CI |
| KMT2D | 0.110 | 0.39 | 0.78-8.29 |
| BCL7A | 0.128 | 2.43 | 0.13-1.34 |
| IRF8 | 0.184 | 1.24E-08 | 0-Inf |
| ARID1A | 0.240 | 1.27E-08 | 0-Inf |
| CREBBP | 0.322 | 1.8 | 0.17-1.81 |
| IGLL5 | 0.393 | 1.66 | 0.19-1.96 |
| BCL2 | 0.481 | 0.63 | 0.44-5.76 |
| STAT6 | 0.486 | 1.7 | 0.13-2.67 |
| TNFRSF14 | 0.496 | 1.47 | 0.92-3.47 |
| FOXO1 | 0.779 | 0.75 | 0.17-10.43 |

**Table S6: Immune genes by maximally selected rank statistics**

| Immune Gene | MaxStatCut Point | High Risk % | High expression associated with high risk | HR | 95% CI | P Value | Adjusted P Value |
|---|---|---|---|---|---|---|---|
| CD3D | 1047.72 | 81.44 | N | 0.57 | 0.27- 1.22 | 0.147 | 0.262 |
| CD4 | 298.11 | 46.39 | Y | 1.30 | 0.78– 2.29 | 0.300 | 0.300 |
| CD8A | 517.7 | 26.80 | N | 0.45 | 0.25– 0.79 | 0.005 | 0.037 |
| FOXP3 | 753.41 | 80.41 | N | 0.66 | 0.31– 1.4 | 0.278 | 0.300 |
| CD68 | 554.33 | 70.10 | Y | 1.76 | 0.92– 3.35 | 0.086 | 0.262 |
| CD163 | 823.16 | 17.53 | Y | 1.58 | 0.81– 3.08 | 0.175 | 0.262 |
| CD47 | 1660.18 | 82.47 | N | 0.60 | 0.28– 1.28 | 0.187 | 0.262 |

**Table S7: Clinical characteristics of Comparator Cohort of Advanced-Stage Follicular Lymphoma Patients (AusASFL)**

| Patient Characteristics | | AusASFL | |
|---|---|---|---|
| | | N | % |
| All Patients | | 68 | |
| Sex | Male | 35/68 | 51.5 |
| Age | ≥60 | 38/68 | 55.9 |
| Stage | 1 | 68/68 | 100.0 |
| FDG-PET Staging | Yes | 68/68 | 100.0 |
| Extranodal Site | Yes | 29/68 | 42.7 |
| Histologic Grade | 1-2 | 10/63 | 15.9 |
| LDH | Raised > ULN | 31/67 | 46.3 |
| Treatment | Watchful waiting | 0/67 | 0.0 |
| | Radiotherapy Alone | 0/67 | 0.0 |
| | R-CHOP | 43/67 | 64.2 |
| | R-Bendamustine | 22/67 | 32.4 |
| | O-Bendamustine | 2/67 | 3.0 |
| Maintenance | CD20 mAb | 49/67 | 73.1 |

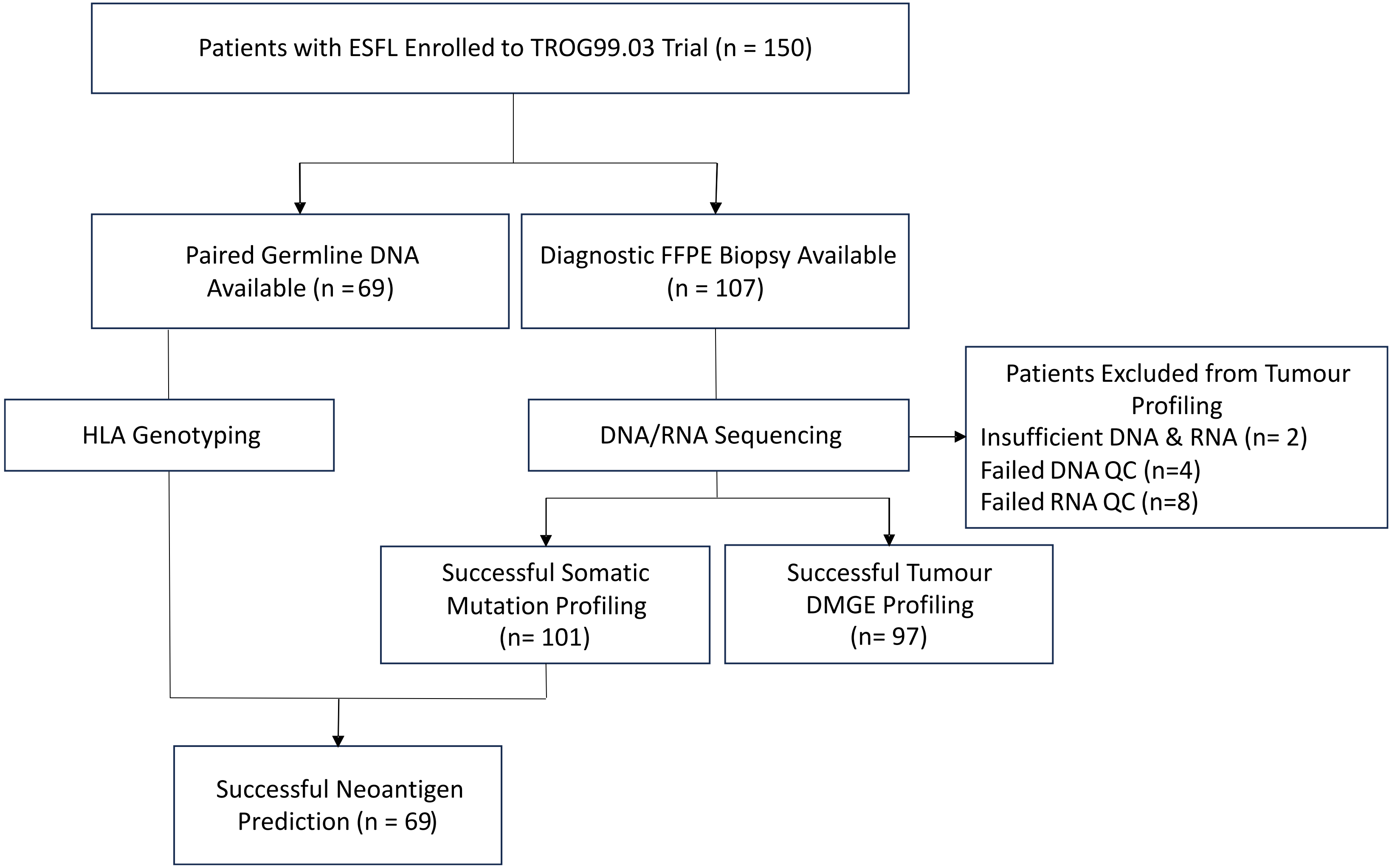


**Figure S1. Consort diagram demonstrating composition of TROG99.03 discovery for translational studies.**

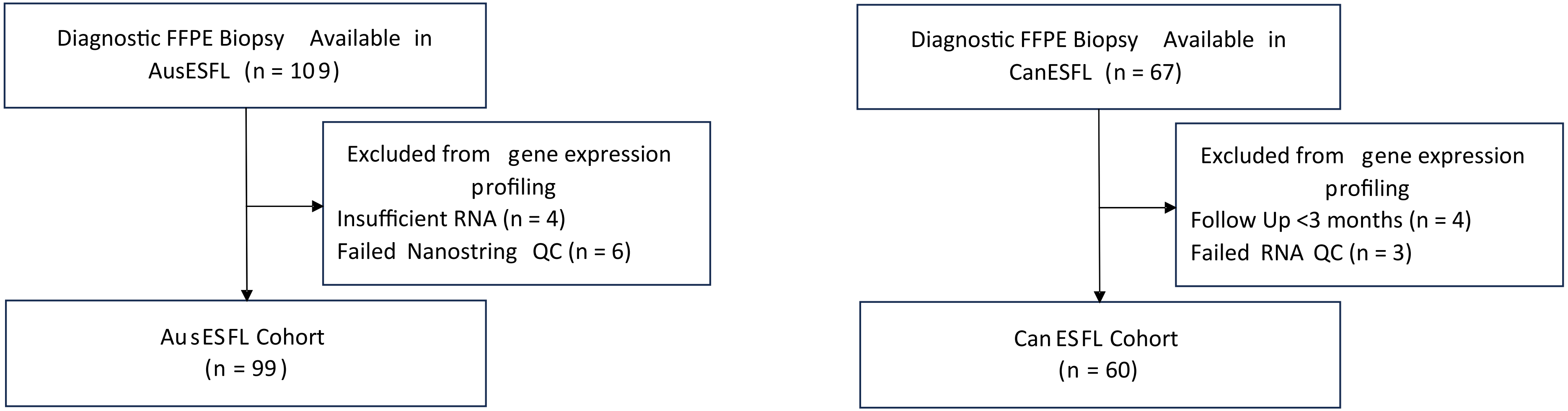


**Figure S2. Consort diagram demonstrating composition of AusESFL validation and CanESFL validation cohorts.**

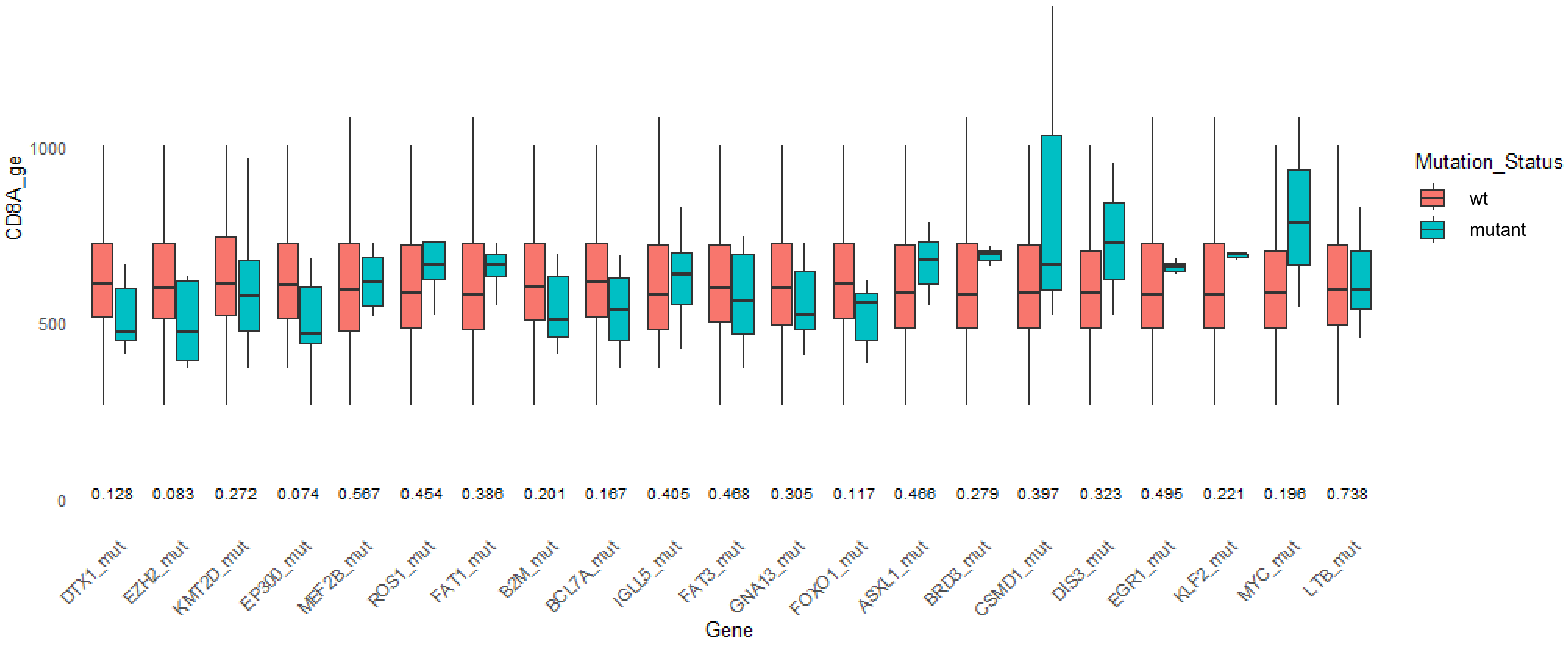


**Figure S3. Relationship between genomic variants and CD8A gene expression.**

Box and whisker plot showing digital multiplex gene expression of CD8A trancriptome. CD8A DMGE is compared by Wilcox rank-sum test to calculate p-values (denoted below the plot) for each gene based on non-synonymous mutation status.
